# Estimated SARS-CoV-2 Seroprevalence in Healthy Children and Those with Chronic Illnesses in The Washington Metropolitan Area as of October 2020

**DOI:** 10.1101/2021.01.30.21250830

**Authors:** Burak Bahar, Joelle N. Simpson, Cara Biddle, Andrew Campbell, Jeffrey S. Dome, Roberta L. DeBiasi, Catriona Mowbray, Stefanie Marguilies, Adrienne Sherman, Jacqueline Reuben, Meghan Delaney

## Abstract

The estimated SARS-CoV-2 seroprevalence in children was found to be 9.46% for the Washington Metropolitan area. Hispanic/Latinx individuals were found to have higher odds of seropositivity. While chronic medical conditions were not associated with having antibodies, previous fever and body aches were predictive symptoms.

## Introduction

Children’s National Hospital (CNH) observed its first pediatric case of severe acute respiratory syndrome coronavirus 2 (SARS-CoV-2) infection in mid-March 2020.^1^ Since then, Coronavirus disease 2019 (COVID-19) has been spreading broadly in the region.^2^ As of mid-January 2021, our institution had performed more than 50,700 SARS-CoV-2 reverse transcriptase-polymerase chain reaction (RT-PCR) tests for the pediatric community. The monthly percent positivity ranged between 2.84% to 11.10% with an average 6.1% viral positive rate. We investigated the prevalence of anti-SARS-CoV-2 antibodies among pediatric patients in the Washington Metropolitan area (District of Colombia and parts of Maryland, Virginia and West Virginia), in the late summer and early autumn of 2020, in collaboration with the District of Columbia Department of Health (DC Health).

## Methods

This study was reviewed and approved by both CNH and DC Health Institutional Review Boards. From July 7^th^ to October 29^th^, 2020, convenience samples of pediatric patients aged 22 years and younger were obtained from CNH Hematology Oncology Clinic, CNH Primary Care Center, two CNH Emergency Departments, and three public, community-based testing sites operated by DC Health. Consenting participants completed a survey that collected information on demographics, history of COVID-19 related symptoms since March 1, 2020 (fever, headache, body ache, fatigue, cough, nausea, vomiting, sore throat, loss of smell, loss of taste and diarrhea), and comorbidities. Blood samples were obtained for serologic testing using standard venipuncture technique. The main outcome was the seroprevalence of anti-SARS-CoV-2 antibodies. The DiaSorin Liaison XL SARS-CoV-2 immunoglobulin G (IgG) S1/S2 assay was utilized for all participants from serum or plasma samples. Seropositivity was qualitatively defined as the presence of anti-SARS-CoV-2 IgG antibodies ≥15 absorbance units per milliliter (AU/mL) according to manufacturer’s threshold.

Age was categorized as 0 through 5, 6 through 15 and 16 through 22 years. Participants self-identified race and ethnicity from fixed categories. Individuals who reported Hispanic/Latinx ethnicity, but reported a different race, were included in the Hispanic/Latinx category. After merging of reported race categories, the race variable consisted of Black, White, Hispanic/Latinx and other (biracial/multiracial, Asian, American Indian, etc.).

Statistical analyses were performed using R software (version 4.0.3). Statistical significance was defined at a *p* value of <.05. Missing data were excluded. Odds ratios and 95% confidence intervals (CI) were calculated with bivariate logistic regression analyses to assess demographic characteristics, symptoms and comorbidities associated with seropositivity. Variables of interest identified in bivariate analyses were included in the multivariate model. The seropositivity rate was adjusted for a test performance of 97.6% sensitivity and 99.3% specificity, as reported by FDA^3^, using the Blaker method^4^ in the epiR package (version 2.0.19).

## Results

From July to October 2020, a total of 385 individuals between 2 months and 22 years old participated, and 38 individuals were found to have antibodies against SARS-CoV-2 (Table 1). After adjustment for test accuracy, the estimated SARS-CoV-2 seroprevalence in the Washington Metropolitan area was found to be 9.46 (95% CI 6.68–13.00) cases per 100 children at risk.

**Table.**
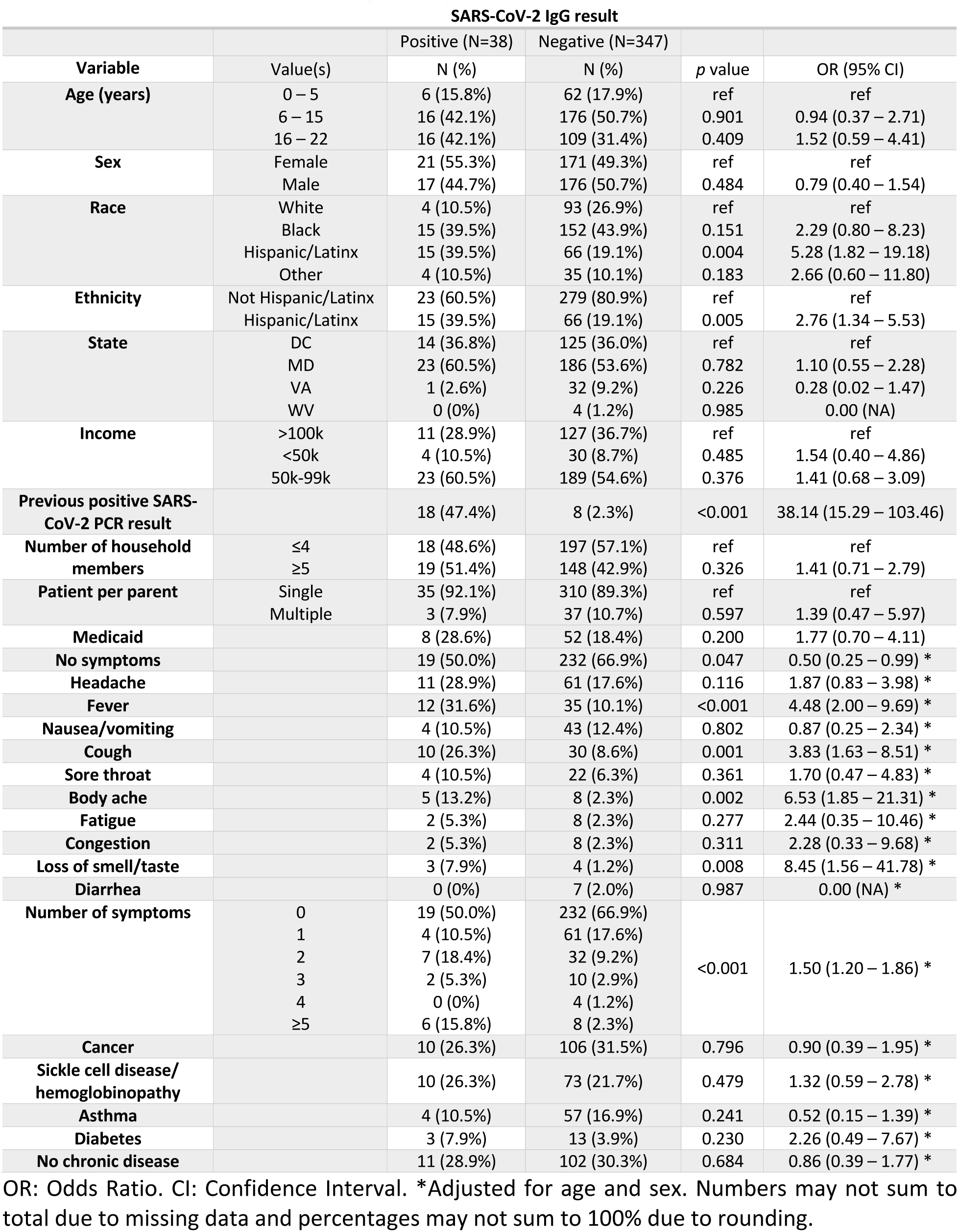
Demographics, comorbidities, previous molecular test and symptomatology as predictors of anti-SARS-CoV-2 antibodies among participants

| SARS-CoV-2 IgG result |  |  |  |  |  |
| --- | --- | --- | --- | --- | --- |
| Variable | Value(s) | Positive (N=38)<br>N (%) | Negative (N=347)<br>N (%) | p value | OR (95% CI) |
| Age (years) | 0 – 5 | 6 (15.8%) | 62 (17.9%) | ref | ref |
|  | 6 – 15 | 16 (42.1%) | 176 (50.7%) | 0.901 | 0.94 (0.37 – 2.71) |
|  | 16 – 22 | 16 (42.1%) | 109 (31.4%) | 0.409 | 1.52 (0.59 – 4.41) |
| Sex | Female | 21 (55.3%) | 171 (49.3%) | ref | ref |
|  | Male | 17 (44.7%) | 176 (50.7%) | 0.484 | 0.79 (0.40 – 1.54) |
| Race | White | 4 (10.5%) | 93 (26.9%) | ref | ref |
|  | Black | 15 (39.5%) | 152 (43.9%) | 0.151 | 2.29 (0.80 – 8.23) |
|  | Hispanic/Latinx | 15 (39.5%) | 66 (19.1%) | 0.004 | 5.28 (1.82 – 19.18) |
|  | Other | 4 (10.5%) | 35 (10.1%) | 0.183 | 2.66 (0.60 – 11.80) |
| Ethnicity | Not Hispanic/Latinx | 23 (60.5%) | 279 (80.9%) | ref | ref |
|  | Hispanic/Latinx | 15 (39.5%) | 66 (19.1%) | 0.005 | 2.76 (1.34 – 5.53) |
| State | DC | 14 (36.8%) | 125 (36.0%) | ref | ref |
|  | MD | 23 (60.5%) | 186 (53.6%) | 0.782 | 1.10 (0.55 – 2.28) |
|  | VA | 1 (2.6%) | 32 (9.2%) | 0.226 | 0.28 (0.02 – 1.47) |
|  | WV | 0 (0%) | 4 (1.2%) | 0.985 | 0.00 (NA) |
| Income | >100k | 11 (28.9%) | 127 (36.7%) | ref | ref |
|  | <50k | 4 (10.5%) | 30 (8.7%) | 0.485 | 1.54 (0.40 – 4.86) |
|  | 50k-99k | 23 (60.5%) | 189 (54.6%) | 0.376 | 1.41 (0.68 – 3.09) |
| Previous positive SARS-CoV-2 PCR result |  | 18 (47.4%) | 8 (2.3%) | <0.001 | 38.14 (15.29 – 103.46) |
| Number of household members | ≤4 | 18 (48.6%) | 197 (57.1%) | ref | ref |
|  | ≥5 | 19 (51.4%) | 148 (42.9%) | 0.326 | 1.41 (0.71 – 2.79) |
| Patient per parent | Single | 35 (92.1%) | 310 (89.3%) | ref | ref |
|  | Multiple | 3 (7.9%) | 37 (10.7%) | 0.597 | 1.39 (0.47 – 5.97) |
| Medicaid |  | 8 (28.6%) | 52 (18.4%) | 0.200 | 1.77 (0.70 – 4.11) |
| No symptoms |  | 19 (50.0%) | 232 (66.9%) | 0.047 | 0.50 (0.25 – 0.99) * |
| Headache |  | 11 (28.9%) | 61 (17.6%) | 0.116 | 1.87 (0.83 – 3.98) * |
| Fever |  | 12 (31.6%) | 35 (10.1%) | <0.001 | 4.48 (2.00 – 9.69) * |
| Nausea/vomiting |  | 4 (10.5%) | 43 (12.4%) | 0.802 | 0.87 (0.25 – 2.34) * |
| Cough |  | 10 (26.3%) | 30 (8.6%) | 0.001 | 3.83 (1.63 – 8.51) * |
| Sore throat |  | 4 (10.5%) | 22 (6.3%) | 0.361 | 1.70 (0.47 – 4.83) * |
| Body ache |  | 5 (13.2%) | 8 (2.3%) | 0.002 | 6.53 (1.85 – 21.31) * |
| Fatigue |  | 2 (5.3%) | 8 (2.3%) | 0.277 | 2.44 (0.35 – 10.46) * |
| Congestion |  | 2 (5.3%) | 8 (2.3%) | 0.311 | 2.28 (0.33 – 9.68) * |
| Loss of smell/taste |  | 3 (7.9%) | 4 (1.2%) | 0.008 | 8.45 (1.56 – 41.78) * |
| Diarrhea |  | 0 (0%) | 7 (2.0%) | 0.987 | 0.00 (NA) * |
| Number of symptoms | 0 | 19 (50.0%) | 232 (66.9%) | <0.001 | 1.50 (1.20 – 1.86) * |
|  | 1 | 4 (10.5%) | 61 (17.6%) |  |  |
|  | 2 | 7 (18.4%) | 32 (9.2%) |  |  |
|  | 3 | 2 (5.3%) | 10 (2.9%) |  |  |
|  | 4 | 0 (0%) | 4 (1.2%) |  |  |
|  | ≥5 | 6 (15.8%) | 8 (2.3%) |  |  |
| Cancer |  | 10 (26.3%) | 106 (31.5%) | 0.796 | 0.90 (0.39 – 1.95) * |
| Sickle cell disease/hemoglobinopathy |  | 10 (26.3%) | 73 (21.7%) | 0.479 | 1.32 (0.59 – 2.78) * |
| Asthma |  | 4 (10.5%) | 57 (16.9%) | 0.241 | 0.52 (0.15 – 1.39) * |
| Diabetes |  | 3 (7.9%) | 13 (3.9%) | 0.230 | 2.26 (0.49 – 7.67) * |
| No chronic disease |  | 11 (28.9%) | 102 (30.3%) | 0.684 | 0.86 (0.39 – 1.77) * |

OR: Odds Ratio. CI: Confidence Interval. \*Adjusted for age and sex. Numbers may not sum to total due to missing data and percentages may not sum to 100% due to rounding.

Age, sex, number of household members, multiple participation from the same family, Medicaid membership, median household income based on reported zip code and state of residency were not found to be associated with having antibodies (*p* > 0.05 [all]); however, both Hispanic/Latinx race and ethnicity were found to be predictors of seropositivity compared to White race (*p*: 0.004; OR [95% CI]: 5.28 [1.82–19.18]) and non-Hispanic/Latinx ethnicity (*p*: 0.005; OR [95% CI]: 2.76 [1.34–5.53]).

Most of the participants (251/385) reported no previous symptoms. Of the individuals who tested positive for anti-SARS-CoV-2 antibody, the most commonly reported symptoms experienced since March 1, 2020 were fever (31.6%), headache (28.9%) and cough (26.3%). After adjustment for age and sex, bivariate logistic regression showed that total number of symptoms (*p*: <0.001; OR [95% CI]: 1.50 [1.20–1.86]), fever (*p* < 0.001; OR [95% CI]: 4.48 [2.00–9.69]), cough (*p*: 0.001; OR [95% CI]: 3.83 [1.63–8.51]), body ache (*p*: 0.002; OR [95% CI]: 6.53 [1.85–21.31]) and loss of smell and/or taste (*p*: 0.008; OR [95% CI]: 8.47 [1.56–41.81]) significantly predicted seropositivity. Fever (*p*: 0.007; OR [95% CI]: 3.19 [1.32–7.23]) and body ache (*p*: 0.071; OR [95% CI]: 3.29 [0.85–11.72]) were the only predictor variables included in the multivariate logistic regression.

Due to our enrollment approach, most of the participants (69.7%) reported a chronic medical condition. However, bivariate logistic regression analysis showed that neither having a chronic medical condition nor the type of chronic medical condition (oncological disease, hematological disease [sickle cell disease or hemoglobinopathy], asthma or diabetes) were predictive of SARS-CoV-2 seropositivity (*p* > 0.05 [all]). While a previous SARS-CoV-2 PCR test result (n=26) was a significant predictor of seropositivity (*p*: <0.001; OR [95% CI]: 38.14 [15.29– 103.46]), 8 children (3 with an oncological, 4 with an hematological and 1 with no chronic medical conditions) with a previous positive SARS-CoV-2 PCR test result did not demonstrate antibodies in their samples. No symptoms were reported by these children and none of the studied variables were predictors of seronegativity (*p* > 0.05 [all]).

## Discussion

The seroprevalence of SARS-CoV-2 among the pediatric population estimated in our analysis of 9.46% was higher than previous studies that included immunocompromised participants or healthy children.^5, 6^ Furthermore, there was no difference in the odds of being seropositive between chronic illness groups, which is particularly notable since immunocompromised children accounted for a large portion of our sample. Given our approach, which included both healthy and individuals with chronic illnesses, and accounted for test accuracy, we believe our estimate is a close approximation of seroprevalence for the diverse pediatric population in our region.

A systematic review of 18 studies on COVID-19 symptomatology in children reported fever and cough to be the most common COVID-19-related symptoms, other symptoms to be present in less than 10-20% of patients in the reported studies, and asymptomatic individuals to range from 14.6% to 42% in this age group.^7^ These findings are similar to our observations. Furthermore, parallel to our previous report of higher rates of SARS-CoV-2 infection in minority children^8^, Hispanic/Latinx children had a higher seropositivity rate compared to whites which was previously reported for adults for the Baltimore-Washington, DC region.^9^ We also observed antibody loss/non-presence in participants with mild symptomatology which is a known phenomenon for COVID-19.^10^

The present work has some limitations. DC Health and CNH sites used similar, but not identical questionnaire instruments. Statistical models were not adjusted for correlated antibody results from the same household; however, this situation only counted for 10.4% (n=40) of the participants and most likely had a negligible effect on the seroprevalence estimation. Further, our sampling approach resulted in the inclusion of more chronically ill children than healthy children, which may introduce selection and reporting biases. Despite these limitations, the current analysis has several strengths. The pediatric sample achieved demonstrated great demographic diversity, and participants enrolled at CNH sites provided consent to be contacted in the future for repeated testing, enabling follow up studies to be carried out. Most notably, children with underlying medical illnesses have not been studied in this way, and as a result, we feel our findings offer important information as all children, whether living with chronic illness or not, must be considered for “back to school” transitions.

Although we report a higher seroprevalence than other studies, our observed 9.46% seroprevalence rate remains well below the levels at which herd immunity has been estimated to occur^11^, however, per CDC’s reports, there is an increased seropositivity rate trend for most of the US states^12^. Future studies should focus on longitudinal seropositivity assessments among children to determine the impact of continued infections in the community, vaccine implementation, and returning to school and extracurricular programs for much needed social, emotional and behavioral development.

## Data Availability

Deidentified data will be provided upon approval by the institutional review boards.

